# Neuroinflammation at the Grey-White Matter Interface in Active-Duty United States Special Operations Forces

**DOI:** 10.1101/2024.05.13.24307070

**Authors:** Brian L. Edlow, Chieh-En J. Tseng, Natalie Gilmore, Isabella R. McKinney, Samantha L. Tromly, Katryna B. Deary, Collin G. Hu, Brian C. Healy, Christine L. Mac Donald, Kristen Dams-O’Connor, Douglas N. Greve, Yelena G. Bodien, Daniel P. Perl, Jacob M. Hooker, Nicole R. Zürcher

## Abstract

Emerging evidence from autopsy studies indicates that interface astroglial scarring (IAS) at the grey-white matter junction is a pathological signature of repeated blast brain injury in military personnel. However, there is currently no neuroimaging test that detects IAS, which is a major barrier to diagnosis, prevention, and treatment. In 27 active-duty United States Special Operations Forces personnel with high levels of cumulative blast exposure, we performed translocator protein (TSPO) positron emission tomography (PET) using [^11^C]PBR28 to detect neuroinflammation at the cortical grey-white matter interface, a neuroanatomic location where IAS has been reported in autopsy studies. TSPO signal in individual Operators was compared to the mean TSPO signal in a control group of 9 healthy civilian volunteers. We identified five Operators with TSPO signal at the cortical grey-white matter interface that was more than two standard deviations above the control mean. Cumulative blast exposure, as measured by the Generalized Blast Exposure Value, did not differ between the five Operators with elevated TSPO signal and the 22 Operators without elevated TSPO signal. While the pathophysiologic link between neuroinflammation and IAS remains uncertain, these observations provide the basis for further investigation into TSPO PET as a potential biomarker of repeated blast brain injury.

**Disclaimer:** The views expressed in this manuscript are entirely those of the authors and do not necessarily reflect the views, policy, or position of the United States Government, Department of Defense, or United States Special Operations Command, or the Uniformed Services University of the Health Sciences.

## Introduction

Repeated blast exposure (RBE) is associated with a broad spectrum of neuropathology in United States (U.S) military personnel.^1, 2^ Human autopsy studies^1, 3^ and an animal model^4^ indicate that interface astroglial scarring (IAS) at the grey-white matter junction is a signature lesion of RBE, but there is currently no diagnostic test to detect IAS in military personnel prior to brain autopsy. The absence of a diagnostic test for IAS undermines efforts to prevent and treat repeated blast brain injury (rBBI).^5^

Translocator protein (TSPO) positron emission tomography (PET) is a potential biomarker for IAS, because TSPO PET detects neuroinflammation that is believed to precede IAS.^6^ In the recently completed ReBlast Pilot study – Long-term Effects of Repeated Blast Exposure in U.S. Special Operations Forces (SOF) Personnel^7^ – we did not observe an association between cumulative blast exposure, as measured by the Generalized Blast Exposure Value (GBEV),^8^ and TSPO signal at the cortical grey-white matter interface. However, the GBEV scores for all 30 SOF personnel in this study ranged from 387,860 to 363,812,869, which is higher than a previously reported threshold (200,000) at which military personnel experience cognitive, psychological, or physical symptoms,^8^ raising the possibility that a statistical association between cumulative blast exposure and TSPO signal may have been masked by the uniformly elevated GBEV scores.

Here, we performed a secondary analysis of the ReBlast Pilot study dataset to determine if TSPO PET can identify individual Operators with increased TSPO signal at the grey-white matter interface – an anatomic region predisposed to IAS. Our goal was to determine whether TSPO PET should be further investigated as a biomarker of IAS, and hence rBBI, in future studies of blast-exposed military personnel.

## Methods

The ReBlast Pilot study protocol was preregistered at ClinicalTrials.gov (NCT05183087), and a detailed description of the study protocol was previously published.^9^ All participants provided written informed consent, in accordance with a protocol approved by the Mass General Brigham Institutional Review Board and the U.S. Special Operations Command Human Research Protections Office. We enrolled 30 active-duty U.S. SOF who met the following inclusion criteria: 1) male; 2) age 25-45 years; 3) active-duty SOF; 4) prior combat deployment; 5) prior combat exposure during deployment, verified by the Combat Exposure Scale;^10^ and 6) exposure to blast overpressure, verified by an interview-based version of the GBEV.^8^ We excluded individuals with: 1) moderate-severe traumatic brain injury; and 2) imaging contraindication.

SOF personnel traveled to the Massachusetts General Hospital Athinoula A. Martinos Center for Biomedical Imaging for two days, during which [^11^C]PBR28 TSPO PET-magnetic resonance imaging (MRI) was performed as part of a multimodal assessment.^9^ [^11^C]PBR28 TSPO data were acquired on a hybrid PET-MRI scanner, the Siemens BrainPET, which uses a head-only PET camera inserted into the bore of a 3 Tesla TIMTrio MRI scanner.^11^ A T1-weighted multi-echo magnetization-prepared rapid gradient-echo (MEMPRAGE)^12^ scan was acquired at 1 mm isotropic resolution with prospective motion correction^13^ for PET attenuation correction and anatomic localization of the TSPO signal.

[^11^C]PBR28 emission data collected 60-90 minutes post-radioligand injection were divided into five-minute frames, reconstructed to standardized uptake value (SUV) images using an MRI-based attenuation map, realigned,^14^ and averaged (SUV_60-90_). The SUV_60-90_ image was then linearly registered to the participant’s MEMPRAGE scan using FreeSurfer’s spmregister,^15^ skull-stripped, and normalized by the whole brain without ventricles^15^ to account for individual differences in global signal (SUVR_60-90_). Three participants with a TSPO genotype that confers low-affinity binding for [^11^C]PBR28 were excluded from the TSPO PET analyses, yielding a sample size of 27 for this analysis. Additional details regarding TSPO tracer synthesis, PET-MRI data acquisition and quality assessment, TSPO genotyping, and PET reconstruction have been previously described.^7^

The civilian control group was acquired as part of a separate study protocol^15^ approved by the Mass General Brigham Institutional Review Board, with written informed consent provided by healthy volunteers. For comparison with the SOF cohort, we identified nine controls who were male and whose age was ≥ 25 years (i.e., the minimum age in the inclusion criteria for the SOF cohort). The mean +/- SD age of the controls was 30.4 +/- 4.5 years. With respect to race, 6 were white, 2 were Asian, and 1 reported more than one race. In terms of ethnicity, two were Hispanic and seven were non-Hispanic. All controls were imaged on the same PET-MRI scanner, and all PET-MRI and T1 MEMPRAGE acquisition, processing, and analysis parameters for the control group were identical to those used for the SOF cohort.

To generate a cortical grey-white matter interface region of interest (ROI), we processed the SOF and control MEMPRAGE datasets using FreeSurfer,^16^ created labels for grey and white matter, and then created a new ROI along the entire cortical grey-white matter interface that included 25% of the deepest thickness of the cortex and an equal thickness of the adjacent overlying white matter. To identify individual Operators with significantly elevated TSPO signal within the grey-white matter interface ROI compared to the control group, we identified individuals with a z-score above 2 (i.e., 2 standard deviations above the control mean). We calculated the proportion with elevated TSPO and an exact binomial 95% confidence interval, and we used the exact binomial test to assess whether the observed proportion of individual Operators with elevated TSPO was different from the expected proportion of Operators with elevated TSPO, under the null hypothesis that Operators were part of the normal distribution derived from the control dataset (null proportion = 0.023). We also assessed for differences in demographics, blast exposure, combat exposure, and blunt head impacts in Operators with and without increased TSPO signal within the cortical grey-white interface ROI. We used Wilcoxon rank sum tests for continuous variables and Fisher’s exact tests for categorical variables.

## Results

Five of 27 Operators had TSPO signal at the cortical grey-white matter interface that was at least two standard deviations above the control mean (Figure). The proportion of Operators with elevated TSPO was 0.185 (5/27), with a 95% confidence interval of [0.063, 0.381], and when we tested whether the observed proportion was different from 0.023, the p-value was 0.0003.

**Figure:**
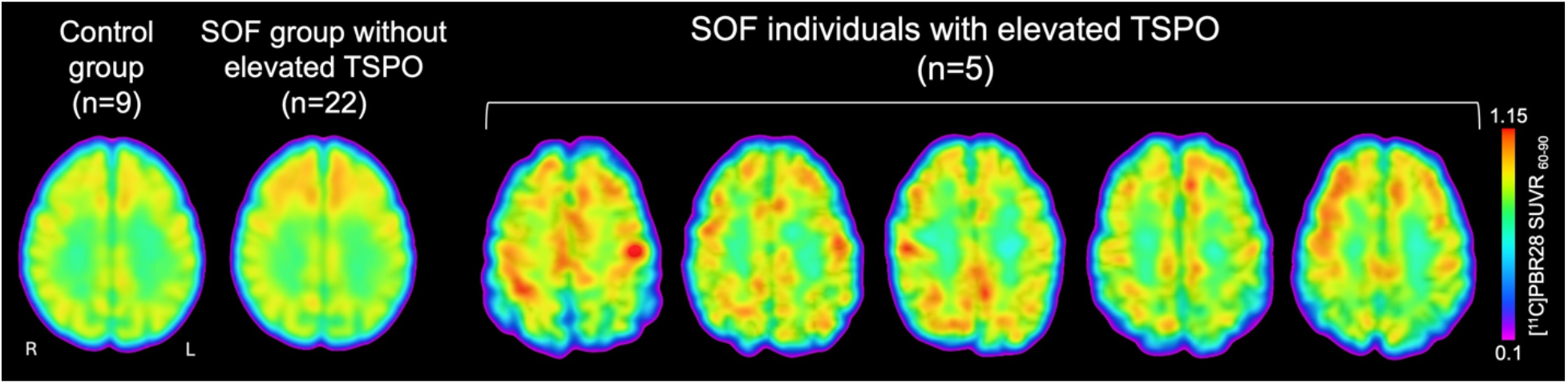
Translocator Protein (TSPO) Positron Emission Tomography (PET) Detects Neuroinflammation at the Cortical Grey-White Matter Interface in Individual Operators. Axial images of TSPO PET data are shown for the five individual Operators with elevated TSPO signal at the cortical grey-white matter interface, compared to the 22 Operators without elevated TSPO signal and to a control group of nine healthy civilians. For the control group and for the 22 Operators without elevated TSPO signal, the mean TSPO PET signal map is shown in MNI standard space. The intensity of the TSPO signal at the cortical grey-white matter interface is indicated by the color bar (bottom inset). Abbreviations: L = left; R = right; SUVR_60-90_ = standardized uptake value normalized by whole brain mean, based on emission data collected from 60-90 minutes post-radioligand injection for [^11^C]PBR28.

The GBEV scores of the five Operators ranged from 389,317 to 38,129,760 with a median value of 19,960,106. Four of five had experienced heavy combat, and one had experienced moderate combat, as measured by the Combat Exposure Scale.^10^ All five had been exposed to explosive blasts within the past year. Four out five had more blunt contact blows to the head than they could recall, as assessed by the Brain Injury Screening Questionnaire.^17^ There were no significant differences in demographics or exposure to blast, blunt head trauma, or combat between the five operators with elevated TSPO signal and the 22 without elevated TSPO signal at the cortical grey-white matter interface (Table).

**Table:** Clinical, Demographic, and Exposure Characteristics. Blows to the head “high” = number of participants with more blows to the head than they could remember; “low” = number of participants who could recall a finite number of blows to the head (range of blows to the head: 1-13), assessed by the Brain Injury Screening Questionnaire (BISQ).^17^ Combat exposure was assessed using the Combat Exposure Scale (CES).^10^ Abbreviations: GBEV = Generalized Blast Exposure Value; NA = not applicable; TSPO = translocator protein. Continuous variables were statistically compared between the two SOF groups using Wilcoxon rank sum tests for continuous variables and Fisher’s exact tests for categorical variables.

| Characteristic | SOF with Non-elevated TSPO (n=22) | SOF with Elevated TSPO (n=5) | P Value |
| --- | --- | --- | --- |
| Age - yr | 36.5 +/- 4.1 | 39.6 +/- 3.4 | 0.34 |
| Sex: Male - no. | 22 | 5 | NA |
| Race: White - no. | 22 | 5 | NA |
| Ethnicity: Non-Hispanic - no. | 19 | 5 | 1.00 |
| Years in Service | 16.5 +/- 4.8 | 19.2 +/-3.0 | 0.47 |
| Military Branch - no. | 15 Army<br>4 Navy<br>2 Air Force<br>1 Marine | 4 Army<br>0 Navy<br>0 Air Force<br>1 Marine | 0.66 |
| Rank - no |  |  |  |
| Officer | 1 | 0 | 1.00 |
| Warrant Officer | 3 | 1 |  |
| Enlisted | 18 | 4 |  |
| CES Score | 33.7 +/- 4.9 | 35.2 +/- 3.4 | 0.90 |
| Combat Exposure (CES) - no. |  |  |  |
| Moderate | 1 | 1 | 0.14 |
| Moderate-Heavy | 9 | 0 |  |
| Heavy | 12 | 4 |  |
| Blows to the Head (BISQ) - no. |  |  |  |
| Low | 8 | 1 | 0.64 |
| High | 14 | 4 |  |
| Cumulative Blast Exposure (GBEV) median [range] | 10,041,994<br>[1,430,877 to 363,812,869] | 19,960,106<br>[389,317 to 38,129,760] | 0.84 |
| Most Recent Blast Exposure - no. |  |  |  |
| <1 year | 18 | 5 | 1.00 |
| 1 year | 2 | 0 |  |
| 2 years | 2 | 0 |  |

## Discussion

In this TSPO PET analysis of active-duty U.S. SOF, TSPO signal at the cortical grey-white matter interface was elevated in five of 27 Operators (18.5%), as compared to a group of healthy civilian controls. There were no significant differences in demographics, blast exposure, blunt head trauma exposure, or combat exposure in individuals with elevated TSPO at the grey-white interface compared to individuals without elevated TSPO at the grey-white matter interface. While the pathophysiological relationship between neuroinflammation and IAS at the grey-white interface remains uncertain, these observations provide the basis for further investigation of TSPO PET as a biomarker of rBBI.

The presence of increased TSPO signal at the grey-white matter interface in active-duty U.S. SOF should be interpreted in the context of evolving concepts about the mechanistic complexity of TSPO signal.^18^ While TSPO has been classically interpreted as a measure of neuroinflammation because of its expression within reactive microglia and astrocytes, there is growing recognition that TSPO may be interpreted in the context of cellular metabolism, given that it is expressed within the mitochondria of neurons, endothelial cells, and vascular smooth muscle cells.^19^ It is therefore possible that changes in TSPO signal may require different mechanistic interpretations, depending on the context.

Our interpretation of increased TSPO signal as a sign of neuroinflammation in the present study is consistent with that of a prior study in a different cohort of blast-exposed SOF personnel.^20^ However, in the previously reported group-level analysis of the same 27 individuals analyzed here,^7^ we found that increased blast exposure was associated with decreased TSPO in the rostral anterior cingulate cortex (rACC), as well as increased cortical thickness at the rACC, and decreased functional connectivity of the executive control network, for which the rACC is a hub node. In this latter context, we interpreted decreased TSPO signal as reflecting mitochondrial-driven metabolic dysfunction within the rACC, particularly because an increase in cortical thickness was interpreted by our group^7^ and others^21^ to be a potential manifestation of IAS.

Collectively, these observations indicate that regional alterations in TSPO signal may reflect distinct pathophysiological processes (i.e., neuroinflammation versus decreased cellular metabolism). It is possible that increased TSPO signal (i.e. neuroinflammation) represents an early, treatable stage of rBBI, whereas decreased TSPO signal (i.e., decreased cellular metabolism) may represent a late stage of rBBI in which IAS has already occurred. Future longitudinal studies of dynamic TSPO signal changes, and their correlation with blast exposure, are needed to clarify the specifics of these potential pathophysiologic mechanisms.

It is also important to consider that pathophysiologic manifestations of neuroinflammation in the human brain are variable and cannot be determined based on an increase in TSPO signal. While there is mechanistic plausibility for a relationship between neuroinflammation and IAS, given the role of reactive astrocytes in IAS pathogenesis, it is also possible that the TSPO signal observed here represents an adaptive neuroinflammatory response to brain injury. Indeed, neuroinflammation may lead to healing of injured neurons and restoration of neuronal function after brain injury.^6^ This potential beneficial role of neuroinflammation is highlighted by the results of a recent clinical trial, in which reduction of neuroinflammation by the anti-inflammatory agent minocycline was associated with increased levels of neurodegenerative proteins in the blood in civilians with traumatic brain injury.^22^ Elucidation of whether neuroinflammation is adaptive or maladaptive in blast-exposed military personnel will require multimodal, longitudinal studies with complementary neuroimaging, blood, cognitive, psychological and pathology endpoints.^23^

In summary, we found evidence of neuroinflammation, as measured by TSPO PET, at the cortical grey-white interface – the same region where IAS has been previously described – in five of 27 active-duty U.S. SOF. Given the small sample size, the use of a civilian control cohort, and the exploratory nature of the analysis, these results should be considered hypothesis-generating. The presence of increased TSPO signal at the grey-white matter interface does not prove a causal link between neuroinflammation and IAS. However, these observations provide the basis for further investigation of TSPO PET as a potential biomarker of neuroinflammation in blast-exposed military personnel. Future studies are needed to clarify which risk factors predispose to neuroinflammation, to determine whether neuroinflammation is a reliable biomarker of rBBI, and to identify predictors of brain healing versus scarring when neuroinflammation is detected in military personnel.

## Data Availability

United States Special Operations Command (USSOCOM) regulations prevent public release of the data generated for the current study. Future requests for these data may be submitted to the corresponding author and will then need to be vetted by USSOCOM.

## Acknowledgments

We thank the SOF personnel who participated in the study. We also acknowledge the following individuals for their contributions to the ReBlast study: John E. Kirsch, Jacob C. Calkins, Amy L. Kendall, and Grae E. Arabasz for serving on the MRI Safety Committee; Mary Tresvalles, Mariah Manter, and Camila Canales, for assistance with PET-MRI data acquisition; nuclear medicine technologists, Shirley Hsu and Oliver Ramsay, for intravenous line placement, radiotracer injection, blood draw, and assistance with PET-MRI scans; the MGH Clinical and Translational Research Unit Core for medical coverage; Anne Siewko and the Martinos Center radiopharmacy team; Alison Brown and Mass General Brigham Personalized Medicine Biobank Genomics Core for genotyping; and Jonathan Hamill, Kevin Kammer, Paul E. Raines, and Sam D. Schoerning for creating the data security systems necessary to conduct the study.

